# Medical brain drain from Western Balkan and Croatia to Germany and Austria – an approach to the digital demography

**DOI:** 10.1101/2021.05.26.21257893

**Authors:** Tado Jurić

## Abstract

**Background:** In this paper, we show that the tools of digital demography, such as the Google Trends (GT) can be very useful for determining, estimating, and predicting the migration of healthcare workers as well as for further predictions of the general interest in emigration.

**Objective:** This paper analyses recent trends in the mobility of health workers in Europe and focuses specifically on the patterns of mobility among medical doctors and nurses from Western Balkans and Croatia to Germany and Austria. It identifies the drivers of this mobility and shows how to predict further migration of this stuff. In the last 10 years, every fourth nurse has emigrated from the Croatian health system -to Germany, while according to projections Croatia will lose as much as 1/3 of all doctors in the next five years.

**Methods:** A special problem in the analysis of the emigration of healthcare workers from the Western Balkans and Croatia is the fact that there is no system for monitoring this process. Official data up to three years late, and exist only for persons who have deregistered from the state system. The basic methodological concept of our approach is to monitor the digital trace of language searches with the Google Trends analytical tool (trends.google.com). Initially, keywords were chosen by brainstorming possible words that we believed to be predictive, specific, and common enough for use in the forecasting of migration of healthcare professionals. To standardize the data, we requested the data for the period from Dec. 2015 to Dec. 2020 and divided the keyword frequency for each word giving us a search frequency index. Then we have compared searches with official statistics to prove the significations of results and correlations.

**Results:** In Croatia search activities using GT for terms such as ‘‘Bewerbung” (job application), “Arbeit” (work), “Krankenschwester, Bewerbung” (nurses, job application) correlate strongly with officially Germans data for emigrants from Croatia. The data collected by this method correlate with official data, which allows reliable forecasts for the future. Austria will soon become one of the most desirable emigration destinations for Croats, especially for healthcare workers. Simultaneously, the emigration of Croatian citizens to Germany will continue.

**Conclusion:** Understanding why health care personnel emigrate from Western Balkans and Croatia and which are the consequences of this process are key to enable state agencies and government to develop optimal intervention strategies to retain medical staff. The benefit of this method is reliable estimates that can enable state agencies and the government to prepare and better respond to a possible shortage of healthcare workers and to protect the functioning of the health system.

## INTRODUCTION

The healthcare system across Europe is facing demographic aging of both staff and users, and this is increasing the demand for health needs and care. The COVID-19 emergency has confirmed that all EU countries have weaknesses in their health system, and one of them is the inadequate supply of health workers. Developed EU countries are tackling the shortage by immigrating health workers, especially from Western Balkans and Croatia. In the countries from which health workers emigrate, negative consequences are felt during the pandemic and the health crisis (Mara, 2020).

A special problem in the analysis of the emigration of healthcare workers from the Western Balkans and Croatia is the fact that there is no system for monitoring this process. Official data up to three years late, and exist only for persons who have deregistered from the state system. This study shows that the analytical tool Google Trends can give useful complement data to the knowledge of demography, especially in the field of migration of healthcare workers.

The population of Croatia today is characterized by declining fertility, natural depopulation, emigration depopulation, and marked aging of the population (Wertheimer-Baletić, 2004a; Nejašmić, 2008a). Undoubtedly, this is a very unfavorable demographic development (Akrap et all. 2017, Wertheimer-Baletić, 2018).

Due to the causality of general development and demographic processes, there is a multiplication of negative consequences in almost all areas of the social life of the Republic of Croatia (Wertheimer-Baletić, 2018). Prolonged depopulation has already had many negative consequences, such as a declining population generation, a reduction in the working population, and an increased need for care for the elderly, while all these unfavorable demographic trends will intensify in the future (Jurić, 2021 a, d).

An additional problem of Croatian society is that due to the decreasing number of children in the family the circle of the main workers of care for the elderly is decreasing, which is a traditional pattern. It will be increasingly difficult for future elderly people to find someone to provide them with immediate care (Podgorelec and Klempić, 2007: 129). After this study from 2007, the situation even worsened (see: trećadob.hr, 2020). The average Croatian pensioner cannot pay for the various types of services he may need from his current income. All this requires the adjustment of the form and type of care for the elderly and the growth of expectations from state care institutions. But in the past 10 years, one in four nurses has moved out of the healthcare system, while the number of those who left directly after completing their education is unknown. At the same time, projections show that Croatia will lose as much as a third of its doctors in the next five years due to emigration and their advanced age (retirement) (Jurić, 2020 a). Meanwhile, the entire neighboring region also faces very similar problems (detailed below).

After briefly showing the results of relevant studies in the next section, we will focus on the emigration of healthcare professionals from Croatia to Germany and Austria and the results we gained with the approach of digital demography. We used Croatia as a case study because there are no studies of this type (digital demography) in Croatia and the wider region of Southeast Europe, and because this region is demographically one of the most affected regions by depopulation in the world.

### Consequences of emigration on the aging population of Croatia, Serbia, and B&H

Low fertility rates are the most important driver of negative population change in Western Balkans and Croatia (UN, 2020). Similar problems with low fertility rates exist also at the EU level (Europa.eu, 2020). However, unlike most EU countries, Croatia, Serbia, and Bosnia and Herzegovina (B&H) have another important problem that is affecting the structure of the population -the emigration of young people.

Since it became a member of the EU in 2013, an average of 50,000 people emigrates from Croatia every year, most often to Germany (85% of all emigrants; Jurić 2017). In the first half of 2019, Croatia, along with Bulgarians, has the highest percentage of emigration of all EU members (EUROSTAT 2019, BAMF, 2019).

Emigration has a direct impact on the pension and workforce system of Croatia, and the same problem affects neighboring countries from which Croatia used to get labor force. The emigration permanently disrupts the biological basis and directly affected the slower population growth and education, pension, and the workforce system throughout the region. Data for B&H shows that in the period from 2014 to 2018, a net reduction in the workforce was 113,000 workers, or 10% of the total workforce (BHAS, 2015-2018; Jurić, Hadžić, 2021). At the same time since 2014, as in the Croatian case, the number of pensioners in B&H has been increasing rapidly (PIO FB&H & PIO RS 2014, 2019). In the period 2014/2015 -2019/2020, the number of pupils in B&H primary schools decreased by 7,76% and in secondary schools by 21,51% (BHAS, 2020 a,b,c). In the period 2014 – 2019, the population of Serbia decreased by 187.688 (RZS, 2020). Similar to in Bosnia and Herzegovina, most permits are issued to citizens in the category 25 – 45 (Jurić, Hadžić, 2021).Of this number, 60,09% emigrated to Germany, 14,16% to Austria, and 4,94% to Slovenia (OECD, 2019). The number of pupils in primary schools is lower by 9.46% compared to the year 2013 and in secondary schools by 7,20% (RZS, 2020, a,b).

According to all available data Bosnia and Herzegovina, Serbia and Croatia will soon run out of potential for economic growth, due to labor emigration (Jurić, Hadžić, 2021).

### Emigration of health workers from the Western Balkans and Croatia to Germany and Austria

The healthcare system across Europe is facing demographic aging of both staff and users, and this is increasing the demand for health needs and care (Arbeitsagentur, 2019). Developed EU countries are tackling the shortage by immigrating health workers. On the other hand, in the countries from which health workers emigrate, negative consequences are felt during the pandemic and the health crisis (see Mara, 2020). This example is perhaps the best illustration of how there is no fair play in the relationship between the center and the periphery of the EU and Europe (Jurić, 2021 a).

Germany and Austria today are undoubtedly addressing its problem of healthcare workers shortages by importing healthcare workers from Western Balkans and Croatia, and this trend becomes even more noticeable during the Coronavirus pandemic (see results). According to official data, every fourth nurse has emigrated from Croatia in the last 10 years -to Germany (see: Jurić, 2020 a; OECD Health Statistics, 2019). Many indices are that the data are very similar for Bosnia and Herzegovina and Serbia (there is no official data after 2017). According to the HSSMS syndicate, Croatia lacked 12,000 nurses even before this epidemic, and according to official statistics, 3,180 nurses and technicians went abroad directly from the system from 2013 to 2018 alone, most often to Germany. In the period from 2009 to 2013, 4,279 nurses emigrated (HZZ – Croatian Employment Service, 2013). The stated data refer only to that medical personnel who were employed in the Croatian health care system, while the number of those who left immediately after finishing school is not known at all. So, in the past ten years alone, 7,500 – 8.000 nurses have left the health care system in Croatia, which makes one in four nurses from the system numbering 28,000 (Jurić, 2020).

According to the Atlas of Croatian Medicine (2020), more than a thousand specialist doctors emigrated, and another 940 are about to leave -they were looking for a letter of resignation. By 2025, the Croatian health care system will have lost another 2,700 doctors due to retirement or emigration, according to the estimates by the Croatian Medical Chamber (2020). When considered that in Croatia in 2020 there were a total of 14,094 doctors, these numbers only become alarming in context.

The average age of emigrated doctors from Croatia is 36 years and the average age of all doctors in the Croatian system is close to 50 (Digital Atlas of Medicine, 2020). When taken into account how many doctors have emigrated or are preparing for this act in 2020 and 2021 (14%), and the fact that in the next five years 2,255 doctors will retire (which is 15% of the total number of employees in health care today), it is to conclude that Croatia will lose more than one-third of all doctors in five years. Primary health care is the most endangered in terms of staff, and there is already a lack of 204 family doctors, 75 pediatricians, and 103 gynecologists. There is a total of 2,142 family doctors in Croatia, but a third of them are over 60 years of age. In Croatian resources, however, no trained staff can replace them (Markovic Huljev, 2020).

On the other side, the German shortage of workers due to the aging population represents an additional concern for the countries of the Western Balkans and Croatia in the context of the workforce. According to projections, the potential labor force in Germany will decrease by 16.2 million workers between 2012 and 2050 for purely demographic reasons (Arbeitsagentur, 2019). The German generations with the highest birth rates will have left working life around 2035. According to model calculations, the net migration with countries of the European Union will soon drop significantly from the current number to slightly below 300,000 (ibid.). For the next 36 years, an average of between 276,000 and 491,000 people would have to immigrate from third countries every year to Germany for the labor force potential to remain constant. It seems to be that the Western Balkans is particularly targeted here. For this reason in 2020 Germany lifted restrictions on the work for people from the Western Balkans (Falkenhain, Raab, 2020; see: Die Westbalkanregelung 2019).

According to a 2015 study by Bertelsmann Stiftung, in the year 2021 Germany must introduce around 100,000 employees in the field of medical care alone, while it is estimated that as many as 200,000 caregivers will be missing to the year 2020 (Bertelsmann Stiftung, 2015). In the EU health sector, an increase of 1.8 million jobs is expected by 2025 (an increase of 8.1% compared to the current situation). In the same period, 50% of health professionals in the European Union are expected to retire or leave the health sector, creating 11.6 million jobs, which is more than in any other sector (Arbeitsagentur 2019). Between 2000 and 2017 employment in the health sector in the EU has been rising by 42%, compared with a 15% rise in overall employment (OECD, 2019). On average, the health sector absorbs about 10% of the workforce (ibid).

According to the German Federal Employment Service, in 2019 over 65,000 citizens of the Western Balkans and Croatia work in the health and care sector in Germany. According to estimates (Faire mobilitaet, 2020), Germany imports more than ten thousand caregivers and nurses from the Western Balkans and Croatia every year, mostly from Bosnia and Herzegovina and Serbia, but also from Croatia, Kosovo, etc. In 2017, of the 4,600 foreign-trained nurses who moved to Germany close to 32% originated from one of the Western Balkans countries (Mara, 2020).

To put these data on 65,288 emigrated health personnel from this region in context, it should be noticed that this is a larger number than the total number of nurses in Croatia and BiH together.

According to the latest data from the German Employment Service, it is evident that every German federal state has a deficit in occupations in the field of care for the elderly, whereas when it comes to the need for nurses there is only one province that does not show a deficit (Bundesagentur für Arbeit, 2020).

Germany also shows a particularly great need in the elderly care sector (Bundesagentur für Arbeit, 2020). It is estimated that in Germany between 300,000 and 600,000 migrant workers are employed in the care of the elderly. These are most often women from Eastern and Southeastern European countries who are mainly mediated to Germany through private agencies. This form of employment is mostly illegal and is becoming more common in Germany and Austria (Freitag, N., 2020). Faire Mobilität (2015) estimates that between 150,000 and 200,000 illegal workers work in the home care sector in Germany alone (Molitor, C., 2015). As German society grows older and faster, so does the number of registered job advertisements in geriatric care has increased 2.5 times in the last ten years (Bundesagentur für Arbeit, 2019:12).

When it comes to doctors, the German official data show that the immigration of foreign doctors, in the amount of ca. 31,000 doctors per year, have successfully filled the gap until 2013 (Institut der Deutschen Wirtschaft Köln, 2014.). The COVID-19 pandemic has made the shortage of doctors obvious again in Germany. According to Deutsches Ärzteblatt (2020), there are numerous cases that German doctors had to continue working even when in contact with people positive for SARS-CoV-2, due to lack of staff (Deutscher Ärzteverlag, 2020). The mismatch between supply and demand will continue to increase during the 2021s, while doctors from the so-called German “baby boom” generation are retiring. This should mean a loss of 20% of all doctors in Germany. Namely, in 2019, 54.1% of all German doctors were older than 65 years (Ibid.).

In Austria, the inflow of foreign-trained doctors has compensated for 60% of outward mobility, but in the case of nurses, the inflow is outpacing the outflow. However, while in 2010 the number of medical graduates was more than two times higher than in the EU28 at 22 per 100,000 inhabitants, over the past decade this ratio has shrunk drastically by one-third to just 14 medical graduates per 100,000 inhabitants – close to the EU28 average (Mara, 2020).

The number of graduate nurses per capita is below the EU28 average and has remained unchanged over the period 2010-2017. Consequently, Austria relies much more on nurses originating from other countries, who accounted for a share of 18% as of 2019, than on medical doctors, with a share at 6% as of 2018 (Mara, 2020). Shortly the demand for health professionals in Austria is expected to surge rapidly. Close to 30% of doctors in Austria are aged 55 and above. According to CEDEFOP, there were more than 13,600 job vacancies for health professionals in Austria at the end of 2019 (Mara, 2020). In the following, we see again how this need for labor has a negative impact on the Western Balkans region. In Serbia and Bosnia and Herzegovina, the loss of health professionals almost doubled between 2010 and 2017, reaching 14% and 8%, respectively, in 2017.

The Western Balkan countries have the lowest density of health professionals relative to their populations while at the same time the number of health graduates in per-capita terms is one of the lowest in Europe (OECD, 2019). Therefore, a further intensification of outward mobility for this category of workers might be devastating for the region (Mara, 2020). Besides, the WHO Global Code of Practice on the International Recruitment of Health Personnel (WHO 2010), deems that recruitment from health systems affected by shortages of health professionals should be avoided.

### Push-and-pull factors of mobility of health workers

The free movement of workers within the EU has had an important impact on mobility patterns, especially for health workers (Glinos, 2015). The phenomenon is very complex, and its drivers are related to economic and institutional factors (Adovor et al., 2019), as well as socio-political factors (Jurić, 2018). On the one hand, the demand and on the other hand the supply and low wages of medical labor are shaping the current situation in the EU. One-third of the EU27 members are affected by the lack of nurses and one-half of the EU28 countries report shortages of medical doctors (OECD Health Statistics, 2019).

Expectations about employment opportunities are recognized as important pull factors for the mobility of healthcare workers. Higher levels of earnings in this sector in a potential host country and relatively high wage differentials between sending and host countries have a positive impact on attracting health professionals in the potential host countries (Mara, 2020).

In 2018 average monthly wages per employee in health work activities in the Western Balkans countries and Croatia were two to three times lower than in the EU28 countries (OECD, 2019).

OECD shows that Health professionals’ wage differential is a pull factor for east-west migration for this category of workers OECD, 2019). The wage differential in the health sector between the EU-CEE countries and Germany and Austria is significant, and as such is an important pull factor of mobility for health professionals from the EU-CEE, Western Balkan countries, and Croatia. Similar patterns and wage gaps apply to nurses (Mara, 2020). Wage differentials in the health sector across the European countries certainly make some of the countries more successful at attracting health professionals than other countries that are failing to retain them (Ibid). Consequently, this group of countries are facing huge challenges to provide health assistance to their own rapidly aging populations, and especially in the context of the COVID-19 crisis.

As a reason for their dissatisfaction and motives for emigration from Croatia, nurses express difficult working conditions due to the insufficient number of employees and non-employment of new nurses, inability to advance in the profession according to education and work experience, many unpaid overtime hours, fatigue, and exhaustion (Koturić-Čabraja, 2020). Besides, the perception of corruption in the country, the feeling of legal inequality, and the general negative social atmosphere that prevailed after the exodus of emigration since 2013 also play an important role in this process (Jurić, 2018).

Another issue that needs to be discussed in this section and this is how the pandemic reflected dissatisfaction with the working conditions of medical staff. The risk of anxiety and other negative mental health reactions among the workforce was described in a viewpoint by Shanafelt et al. (2020). The toll of the crisis has been heavy on health-care workers (Pfefferbaum, North, 2020). During COVID-19, a higher level of occurrence was found for all measured negative personal symptoms and negative professional symptoms (Vanhaecht et al., 2020). The founded association between COVID-19 and mental health was generally the strongest for the age group 30–49 years, females, nurses, and residential care centers (Vanhaecht et al., 2020). Prolonged stress at work can lead to burnout syndrome (Maslach et all, 2001). It is associated with different consequences such as psychosomatic problems, lower employee performance, and greater depression and drug consumption (LLorca-Rubio, 2017). Teachers, police officers, nurses, and doctors have a prevalence in the population between 35% and 40% (Moukarzel et al., 2019).

Although there are still no studies on this issue in Croatia and Western Balkan countries, there are numerous testimonies of nurses and doctors in the Croatian media about dissatisfaction with working conditions during the pandemic, which were manifested in several strikes during the pandemic Novi list, 16.02.2021). According to testimonies, this crisis also contributed to the search for better working conditions through emigration.

## II. METHODS

### Google Trends as a source of data for predicting the migration of healthcare workers

Understanding why health care personnel emigrate from Western Balkans and Croatia and which are the consequences of this process are key to enable state agencies and government to develop optimal intervention strategies to retain this staff and protect the functioning of the health system. A special problem in the whole analysis of the emigration of healthcare workers from the Western Balkans and Croatia is the fact that there is no system for monitoring this process. Official data exist only for persons who have deregistered from the state system, while for nurses who have not been employed, there is no official data. For this purpose, we created a method that can be useful for monitoring this process as well as for further predictions of the general interest in emigration.

Digital footprint monitoring is the major source of innovation in the context of digital demography (see: Zagheni and Weber, 2012; State et al., 2013; Zagheni et al., 2014, 2017, 2018, Dubois et al., 2015, Spyratos et al., 2018). Namely, as more and more people are leaving their digital traces on the web, the use of these data for different types of research is becoming more common. In recent years, great efforts have been made to devise new methods that, in addition to the existing ones, can provide some answers to open questions in the field of demography and public health (see: Dubois, Zagheni, Garimella, Weber 2018 and European Commission 2016).

Every year, the global spread of the Internet and digital technologies radically transforms the way people communicate with each other, and with the advent of the Covid-19 virus that process has been further accelerated. The Covid-19 outbreak and lockdown accelerated the adoption of digital solutions at an unprecedented pace, creating unforeseen opportunities for scaling up alternative approaches to social science (Hantrais et al, 2020). As a result of the fourth digital revolution and the pandemic, individuals have begun to leave an increasing number of traces online.

As the Internet penetration rate accelerates and increases significantly compared to the creation of credible registration systems for monitoring migration and changes in public health, developing tools that retrieve alternative new sources of information is very likely to become an accepted additional approach to monitoring demographic trends of all kinds. Web-assisted research is being demonstrated daily in many other areas such as epidemiology, economics, statistics, and sociology (Spyratos et al., 2018).

By mid-2020, 58% of the world population was estimated to be internet users, compared to almost 90% in the European Union (Internet World Stats, 2020). Within the EU, the same study showed that usage ranged from nearly 98% in Denmark to less than 70% in Bulgaria -in Croatia (81%) (Eurostat, 2020). Eurostat data show that 19 percent of citizens in Croatia have never used the Internet, while the EU average is 11 percent (Eurostat, 2020). The data show that when it comes to the use of Internet services, Croatia is generally comparable to the EU average. As the age increases, so does the percentage of citizens without any experience in using the Internet. That is most people who do not use the Internet come from the group over the age of 65, as many as 62 percent of them (Jurić, 2021 b).

The pandemic accelerated the uptake of digital solutions in data collection techniques (Sogomonjan, 2020). Nevertheless, to our knowledge, research on the use of large data sources (big data) in the field of migration in southeastern Europe does not exist. We certainly believe that the existing gap between “digital” and “classic” demographics can be bridged and that the data in these databases can be supplemented. It is on this track and for this reason that we join the development of new innovative methods based on the concept of “big data” with a focus on migration monitoring of healthcare workers, as well as their predictions. To this end, we will try to set some outlines for model development in this paper. We believe that GT is an extremely useful tool, which, although primarily intended for use in marketing and business purposes to impose consumer preferences, can also be used for scientific research purposes.

### Innovative data sources in demography

Internet data, i.e., digital traces, could become transformative for demography, especially in the field of migration studies. Data obtained by tracking digital traces, in addition to being extremely robust, are easily collected and generated in real-time and provide a very deep insight into the opinions of individuals (Holbrook and Krosnick 2010). Digital traces thus provide a much deeper insight into attitudes, which can be easily disguised in other forms of data collection (Marwick and Boyd 2010). Moreover, digital traces provide documentation of both movement and activities, which can help researchers bypass possible sources of error in survey data, such as inability to recall, bias, and the like. Finally, the use of digital traces can provide access to groups that are difficult to reach or that are generally underrepresented by traditional research techniques (Cesare, 2018: 1983).

A recent report prepared for the European Commission examined the feasibility of using large data to study demographic issues (European Commission et al. 2016) and concluded similarly to the UN that a) large data sources do not replace traditional data sources but can complement them, b) they can still be used to assess trends (Spyratos et all, 2018). However, these data are also characterized by several shortcomings as well as data from classic demographic sources. In this context, we will mention for now only the fact that although traditional sources of migration data are deficient, they are the only ones that can verify the data we obtain from digital sources, so this model by its nature is closely based on classical data sources. We certainly believe that the combination of both is still the way to get more precise insights. In the continuation of the work, we will show that Google Trends can be used to gather insights that have the potential to capture indices for migration planning. They may capture insights into what was going on in the mind of the user through a non-invasive manner (See: Saeb et al., 2016, Ghandeharioun et al., 2017, Wang et al., 2018). The basic hypothesis of this paper is that the analytical tool Google Trends is a useful source of data for determining, estimating, and predicting migrations, in this specific case of healthcare workers from Croatia.

The basic methodological concept of our approach is to monitor the digital trace of language searches with the Google Trends (GT) analytical tool (trends.google.com). The GT analytics application is a trend search tool that shows the popularity of a term when searching on Google, and we can see if a trend is rising or falling. GT does not provide information on the actual number of keyword searches. Instead, it standardizes search volume on a scale of 0 to 100 over the period being examined, with higher values indicating the time when the search volume was greatest, allowing for verifiable metrics (trends.google.com). It should be borne in mind, however, that each of these searches was conducted for its reason and does not answer researchers’ questions, so “googling” the term “Germany” is not necessarily an implication that someone wants to move to Germany, but may be interested in living conditions. tourist information or just looking for the German Bundesliga (Jurić, T., 2021 c, d). Nevertheless, such aggregated data can also be useful for shedding light on certain research questions, which we will try to show later in the paper.

The Google search Index cannot estimate the exact number of searches, so with the help of this tool the exact number of emigrants cannot be estimated, but the increase of the trend can be noticed very precisely. We tested the method in the Croatia case by comparing the findings obtained with GT with official indicators. We show that the increase in Google search for therms in german in Croatia: “Krankenschwester/Krankenpfleger + Bewerbung (Nurse, application for job Germany, Austria), „Anmeldung” (Registration of residence in Germany, Registration of residence in Austria) and „Arbeit in Deutschland”, Arbeit in Österreich” (work in Austria, Germany) is correlated with the increase in emigration recorded by official statistics and that the decrease in Google search is correlated with the decrease in the number of emigrated cases recorded by official statistics.

To standardize the data, we requested the data for the period from Dec. 2015 to Dec. 2020. We then divided the keyword frequency for selected words giving us a search frequency index. We have then compared searches with official statistics to prove the significations of results (see further explanations by Wilde et al., 2020).

Initially, keywords were chosen by brainstorming possible words that we believed to be predictive, specific, and common enough for use in the forecasting of migration. We used a similar method in predicting migration from Croatia (see Jurić, 2020, Digital demography). After the Significance screen, we selected followed keywords and topics.

To understand these terms, a note on the logic behind the Google Trends search algorithm is necessary (Wilde et al., 2020). Certain delimiters, such as “, -, and + allow users to change the combinations of keywords searched. A search for a single keyword will yield the search frequency index counting all searches containing that keyword, including searches that contain other words (Wilde et al., 2020).

### Limitations of the methodological concept

The study we present, like all others of this type, has important limitations that we want to highlight. Although previous research in this area has shown the feasibility of using digital data for demography, at the same time we highlight the problems associated with assessments and conclusions (see: Zagheni, Weber, Gummadi, 2017; Zhang 2020). Namely, it is unquestionable that there are still significant open methodological issues and the questionable integrity of the data obtained using the sources of large data sets (Jurić, 2020 c).

1. Although the data obtained on social networks and the web offer a new number of new opportunities for demography in general, and even though these are robust data with large samples, which provide information qualitatively different from what can be obtained from the CBS report, Eurostat and other official databases, they are still not representative of the observed population.
2. Furthermore, the lack of control over the way data is obtained and processed causes some uncertainty regarding the possibility of using such data effectively for demographic surveys.
3. A special problem exists in the education of the researchers themselves who must be skilled in programming and computational methods, be transparent about their methods to ensure repeatability, and be accustomed to the interdisciplinary environment.
4. From an ethical point of view, it is also questionable how ethical it is to use data that were not collected with the consent of the user, even when personal data is not known because users did not use these applications knowing that they will be studied (Jurić, 2021 d). We want to emphasize that none of the queries in this project’s database can be associated with a particular individual. This project’s database retains no information about the identity, IP address, or specific physical location of any user.
5. A particular limitation of this approach is the fact that the demographic characteristics of users cannot be determined.
6. The last item is both a limitation and an advantage of this approach. Namely, in a traditional research process, a researcher with a pre-defined theoretical framework and questions collects data from a survey using a carefully crafted set of definitions for each item in the survey. With digital data, the reverse process of operationalizing the research concept occurs. The researcher first observes all the activities and then puts them in a theoretical framework. (Cesare et al., 2018: 1983).

Due to all the above mentioned, it is unquestionable that this model has unresolved issues related to the reproducibility of the findings and the validity of the measurements, which arise from the very characteristics of the Google Trends (GT) system used.

Although these open-ended issues pose serious challenges for making clear estimates, statistics offer a range of tools available to deal with imperfect data as well as to develop controls that take data quality into account (see: Zagheni, Weber, Gummadi, 2017; see: “R”; see: Jurić 2021 d).

## RESULTS AND DISCUSSION

### Use of the Google Trends analytical tool to forecasting the migration of health workers

The Google search Index cannot estimate the exact number of searches, so with the help of this tool the exact number of emigrated health care workers cannot be estimated, but the increase of the trend can be noticed very precisely, which can serve as an indicator.

In the context of predicting migration, it is especially interesting to look at the example of Austria. The very fact that in Croatia in 2020 the German term “Lebenslauf” (germ. resume, CV) has been searched for ever more intensively since Croatia acceded to the EU, and that this trend has remained constant over a long period, suggests that many citizens are getting ready to emigrate.

Searching for job applications in Croatia during 2020 (germ. “Bewerbung”) was more common than the search for the equivalent in Croatian (Croat. “zamolba za posao”). It is also evident that this is an upward trend. This is a strong indication that Croatian citizens are preparing to emigrate more and more to Austria and Germany (Jurić, 2021 c).

In Germany and Austria, “application for a job” is the most searchable term in all in the Croatian language, followed by the term CV (Croat. “životopis”), i.e. how to write a CV according to German standards (Jurić, 2021 c).

According to our findings, Austria will soon become one of the most desirable emigration destinations for Croats, especially for health workers. This was revealed through following a group of Croatian terms searched to the category of terms “Austria”, which indicated an increase in the interest in life, housing, and jobs in Austria. Similarly, we found further indicative sets for the entries such as “registration of residence in Austria” and “deregistration of residence from Croatia”. We checked these findings also from an Austrian perspective.

A further indication is a search for terms related to the registration of residence in Austria and Germany in combination with the entry “PCR Test”. Since the entry “PCR Test” was unknown in Croatia before the outbreak of the pandemic, we believe that this finding is a strong signpost on how to monitor the current situation.

**Figure 1:**
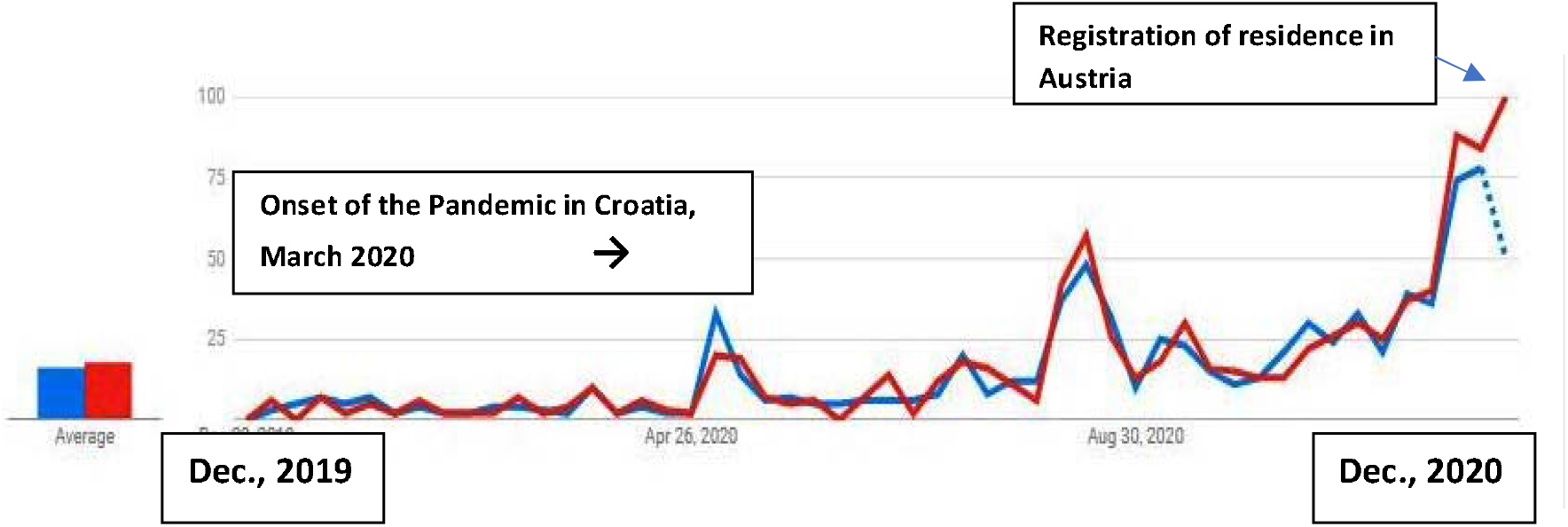
Search queries in German from Croatia “Registration of residence in Germany + PCR; Registration of residence in Austria + PCR Test” (searching for a term in Croatian and German from Croatia) (2019 – 2020)

The Increase in Google search queries “Registration of residence in Austria” follows a similar pattern that we observed in the case of migration of Croatian citizens to Germany in the year 2014 (Jurić, 2021 c). We noticed that the search for the term “Registration of residence in Germany” was particularly intensive before Croatia acceded to the EU (2013) and in the year following the announcement of the lifting of the employment restriction (Jurić, 2021 c). We took this finding as an initial assumption that the migration of Croatian citizens can be predicted. Migrations are most intense in the year following the lifting of employment restrictions in EU states. We compared this prediction with the findings from Austria, Sweden, and Ireland, all of which confirmed the same matrix.

**Figure 2:**
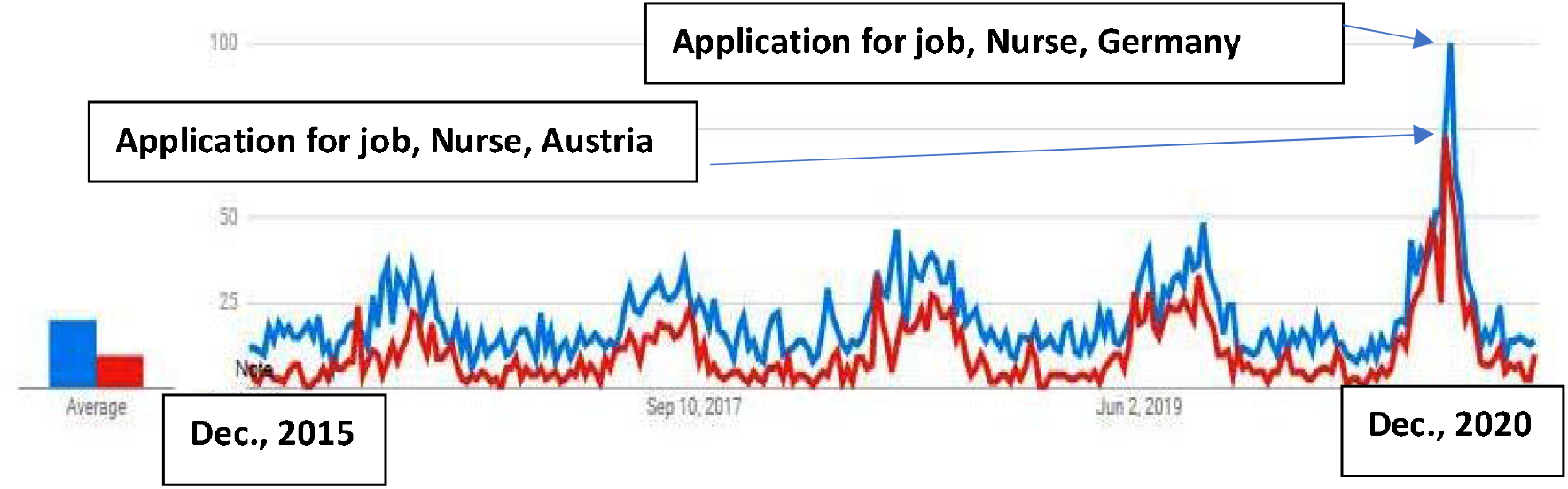
Search queries in German from Croatia „Krankenschwester/Krankenpfleger + Bewerbung Deutschland” and „Krankenschwester/Krankenpfleger + Bewerbung + Austria” (Nurse + application for job Germany + application for job Austria) (2015 – 2020)

Search queries in German from Croatia „Krankenschwester/Krankenpfleger + Bewerbung Deutschland” and „Krankenschwester/Krankenpfleger + Bewerbung + Austria” (Nurse + application for job Germany + application for job Austria) (2015 – 2020) have risen particularly during the pandemic 2020 year. This could be correlated with the difficult working conditions of healthcare workers in Croatian hospitals (Digitalni atlas, 2020), but also with the increased recruitment of healthcare workers from Germany.

German official policy makes no secret of the fact that the import of healthcare workers is a matter of prior national importance. Thus, at the end of 2019, a state agency was opened in Saarbrücken to assist in the transfer of carers and doctors (Deutsche Fachkräfteagentur für Gesundheits-und Pflegeberufe, DeFA). This state agency intends to speed up immigration procedures with the German authorities for healthcare workers recruited by private employment companies, hospitals, and nursing homes from abroad (German Chamber of Industry, 2019). The goal is for foreign nurses to be able to immigrate to Germany within three months of applying for a visa, a process that lasted up to two years until 2019.

**Figure 3:**
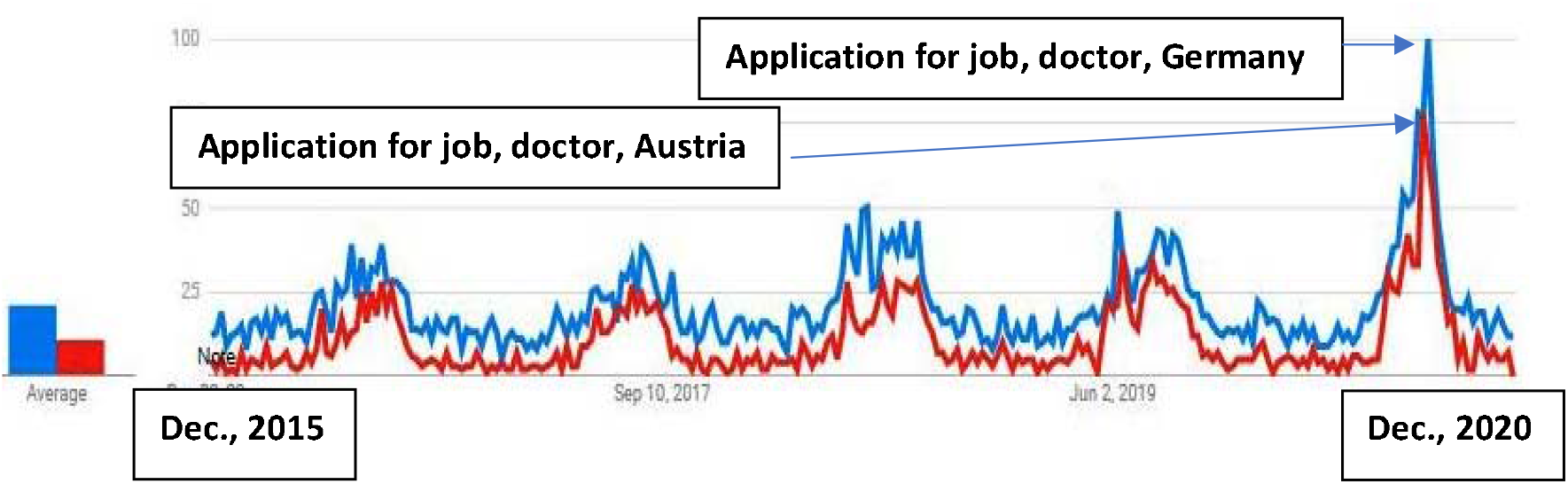
Search queries in German from Croatia „Arzt + Bewerbung Deutschland” and “Arzt + Bewerbung + Austria “(Doctor + application for job Germany + application for job Austria) (2015 – 2020)

All those findings show the growing trend of interest of healthcare workers (doctors, and nurses) to emigrate to Germany and Austria (see: further proceedings).

In further proceedings to standardize the data, we requested the data for the period from Dec. 2015 to Dec. 2020 and divided the keyword frequency for each word (see table 1) giving us a search frequency index. Then we have compared searches with official statistics to prove the significations of results (see BAMF, 2020).

**Table 1:** Emigration from Croatia to Germany shown in a chosen year

| (2012) | (2017) | (2018) | (2019) | (first half of 2020) |
| --- | --- | --- | --- | --- |
| 9,019 | 50,283 | 48,618 | 40,151 | 12,858 |

**Source:** (BAMF 2020)

**Table 2.** Number of nurses from the Western Balkans and Croatia in Germany in 2018 and 2019.

| Year | B&H | Serbia | Kosovo | Albania | North Macedonia | Montenegro | Croatia |
| --- | --- | --- | --- | --- | --- | --- | --- |
| **2018** | 19,631 | 13,598 | 7,204 | 5,206 | 4,094 | 993 | unknown |
| **2019** | 21,789 | 15,216 | 8,050 | 6,642 | 4,487 | 1,104 | from 2011 to 2019: 7,500 |
| **Total: 57,288 from the Western Balkans + 7,500 from Croatia** |  |  |  |  |  |  |  |

Source: Own elaboration from data Arbeitsagentur 2019; Data for Croatia: T. Jurić (2020)

**Table 3.** Medical brain drain (ratio of doctors abroad over the total number of doctors at home and abroad)

| Country | 2010 | 2017 (latest year available) |
| --- | --- | --- |
| Bosnia and Herzegovina | 8% | 14% |
| Serbia | 4% | 8% |

Source: own calculation, UNESCO STATS.

**Table 4.** The stock of doctors emigrated to OECD country’s (mostly to Germany)

| Country | 2010 – 2017 (latest year available) |
| --- | --- |
| Bosnia and Herzegovina | 1,688 |
| Serbia | 3, 339 |

Source: own calculation, using OECD, UNESCO, WHO, and national statistics.

**Table 4.**
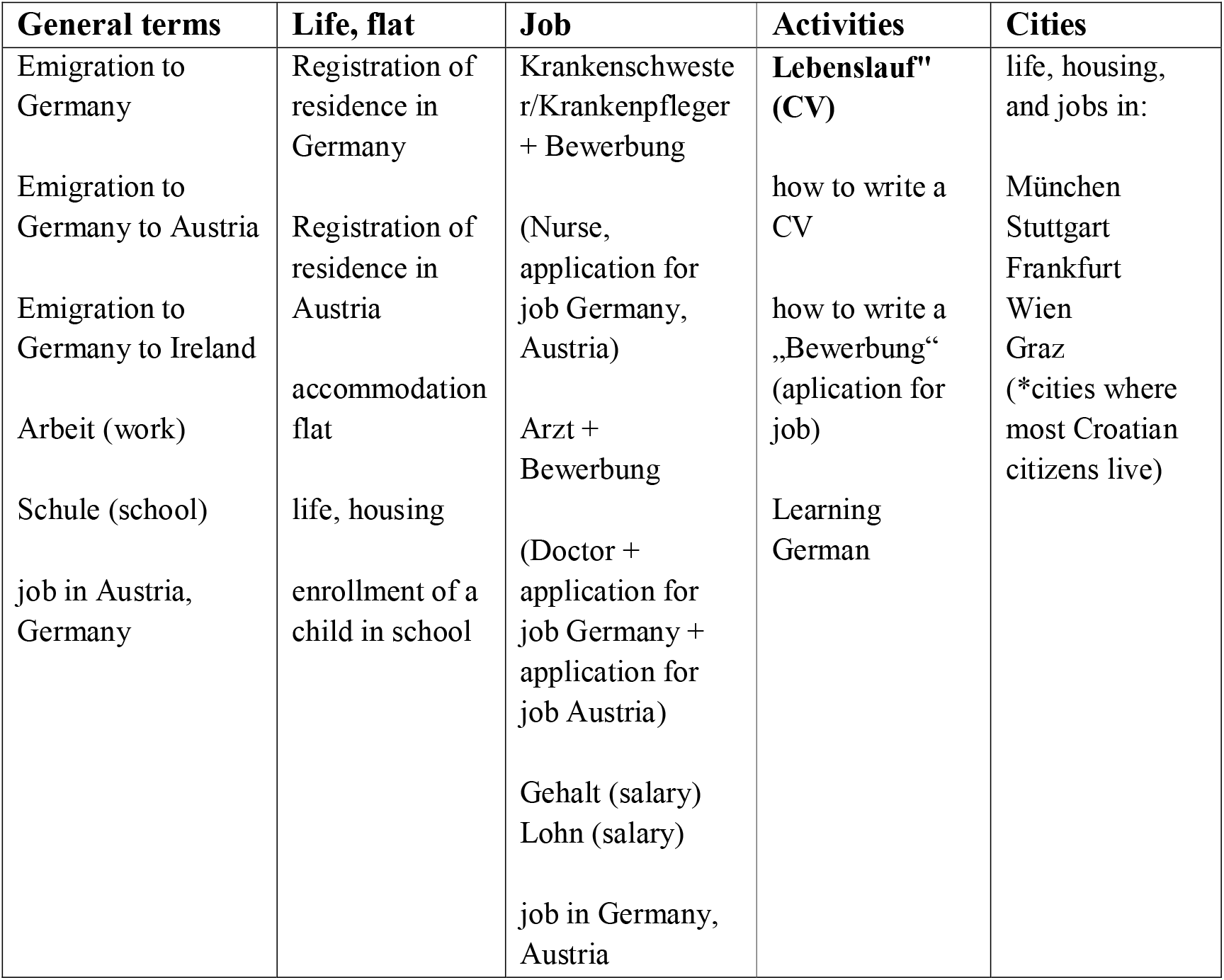
Keyword and topic selection criteria.

**Table 5.** Immigrants from Croatia in Germany 2010 – 2020

| 2010 | 2011 | 2012 | 2013. | 2014. | 2015. | 2016. | 2017. | 2018. | 2019 | *2020<br>(Until<br>June) |
| --- | --- | --- | --- | --- | --- | --- | --- | --- | --- | --- |
| 4836 | 8089 | 9019 | 18.633 | 37.060 | 50.646 | 51.163 | 50.283 | 48.618 | 40.151 | 12.858 |

Source: BAMF, Bundesamt für Migration und Flüchtlinge, Forschungszentrum Migration, Integration und Asyl, Nürnberg, 2017,2018, 2019, 2020\*

**Figure 4.**
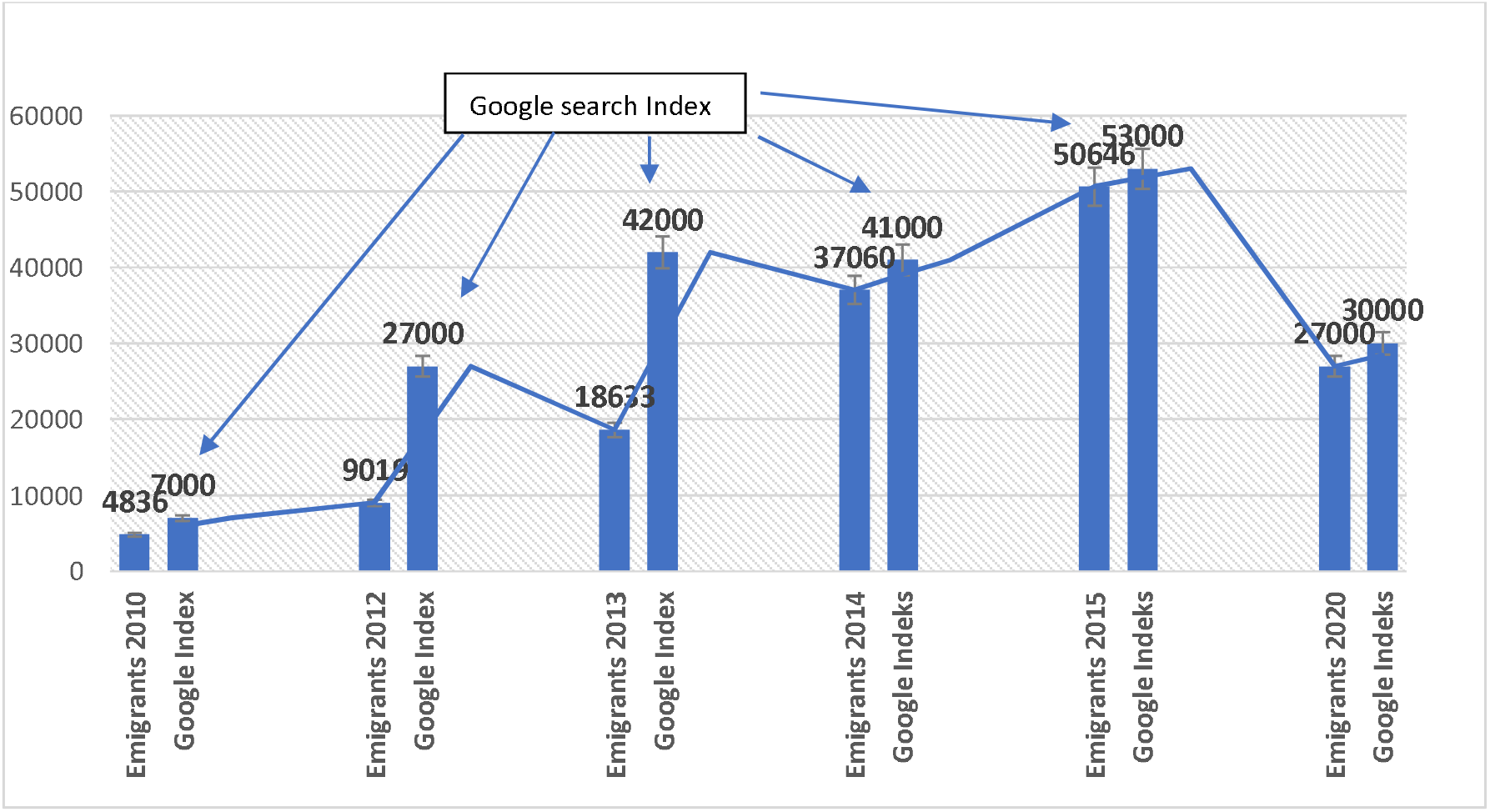
Correlation between Google search Index for query “Arbeit in Deutschland” (work in Germany) in Croatia and the official German statistics for immigrated Croatian citizens by selected years 2010, 2012, 2013, 2014, 2015, 2020

Data for 2020 are only until June because, at the time of submission of this article, data for the whole of 2020 have not yet been published. The data for Google Index is complete. This information should therefore be checked after publication. Here it is important to note that the Google index has not dropped significantly compared to the previous year which is very likely an indication that interest in emigration has not declined, but potential emigrants are waiting for easing movement restrictions so they can emigrate.

**Figure 5.**
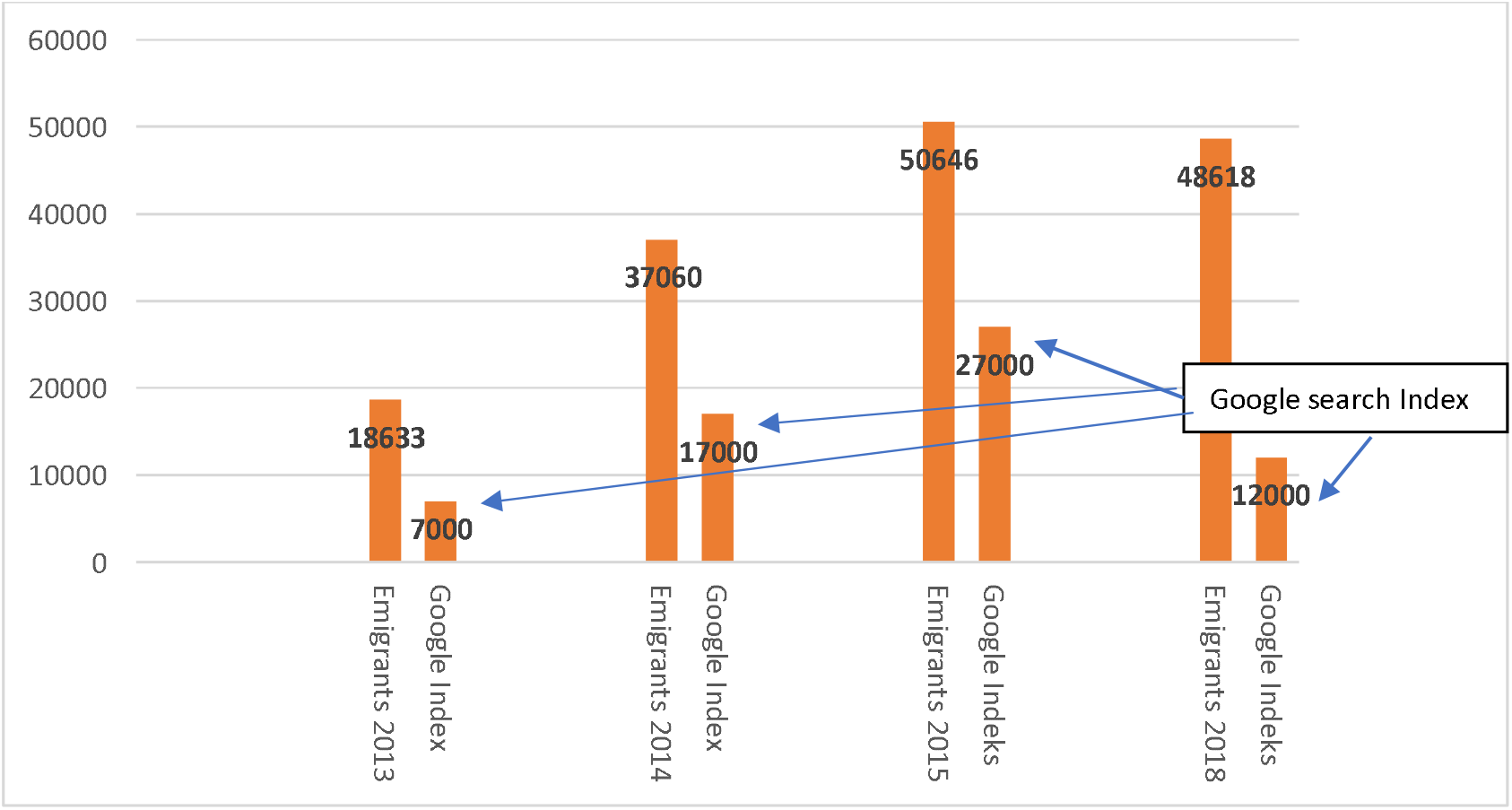
Correlation between Google search Index for query “arbeiten in Deutschland” (working in Germany) in Croatia and the official German statistics for immigrated Croatian citizens by selected years 2010, 2012, 2015, 2019, 2020

**Figure 6.**
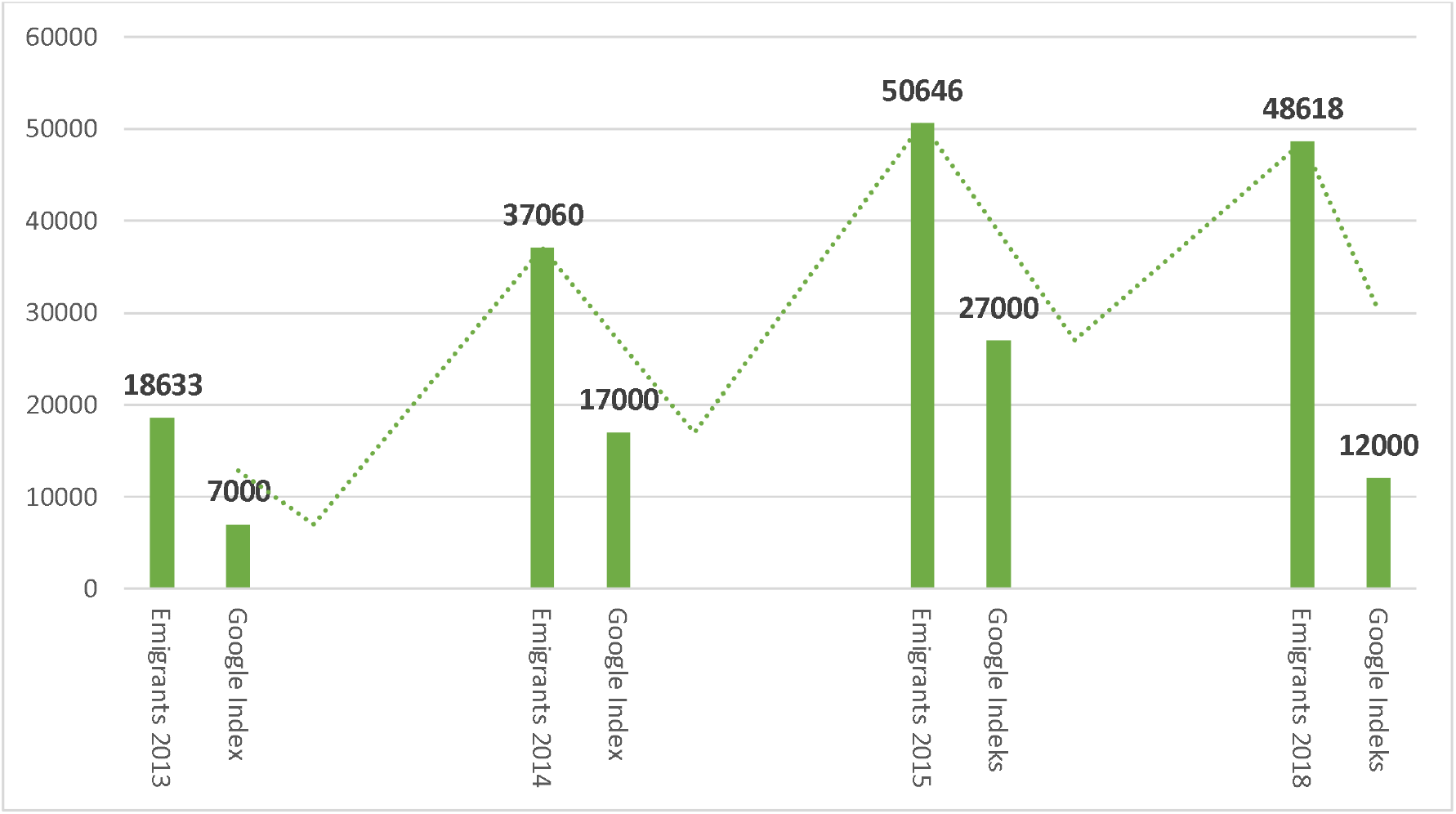
Correlation between Google search Index for query “Bewerbung” (job application) in Croatia and the official German statistics for immigrated Croatian citizens by selected years 2013, 2014, 2015, 2018.

The obtained data also match the results of the survey from Koturić-Čabraja research (2020). This survey showed that the number of retraining in health care occupations has since 2013 increased in Croatia. The survey was conducted at five Zagreb high schools that organize and implement programs for the education of persons over the age of 18-All high schools show that the number of candidates has increased by 50% since 2013 and especially during 2015-2017, with a slight decline in the number of candidates in 2019 and 2020. The largest share is made up of caregivers, gerontological nurses, and nannies, whose main goal is to be educated for working abroad (Koturić-Čabraja, 2020).

All tested correlations show that the increase in Google search, i.e. Google search Index is correlated with the increase of emigration from Croatia and that the decrease in Google search is correlated with the decrease of emigration from Croatia. On the other hand, the increase in total emigration is correlated with the increase in the emigration of medical staff.

The presented method contributes in a way that proves the feasibility of predicting further migrations from Croatia, in this specific case of healthcare workers to Germany and Austria. This tested method shows that the increase in Google search index is correlated with the increase of emigration from Croatia and that the decrease in the Google search index is correlated with the decrease of emigration from Croatia. On the other hand, the increase in total emigration is correlated with the increase in the emigration of medical staff. This allows reliable forecasts for the future.

The insights are particularly relevant for national and EU policymakers and can help design appropriate strategies to retain healthcare workers. The method can enable state agencies and the government to prepare and better respond to the shortage of healthcare workers in the future and protect the functioning of the health system.

## Data Availability

In our work, we use only anonymous, aggregate data. All data are collected following the applicable GDPR and ethical principles of personal data handling.

